# Planning and developing an intervention to improve the psychological wellbeing of people living with persistent pain: De-Stress *Pain*

**DOI:** 10.1101/2024.09.27.24314271

**Authors:** Stephanie Hughes, Tamar Pincus, Adam W A Geraghty, Carolyn A. Chew-Graham, Beth Stuart, Paul Little, Michael Moore, Hollie Birkinshaw

## Abstract

**Background:** People with persistent musculoskeletal (MSK) pain often report depressive symptoms. Distress and depression have been found to predict the development of chronic pain conditions. Evidence suggests pain-related distress is qualitatively different from depressive illness, and current referral pathways and available interventions are sub-optimal for people with persistent MSK pain and distress. We aimed to develop and test the acceptability and proof of concept of an intervention to reduce pain-related distress in people with chronic MSK pain.

**Methods:** The Person-Based Approach (PBA)(1) informed the processes undertaken to inform intervention development. This included semi-structured interviews with people with persistent pain and with General Practitioners (GPs); and a stakeholder discussion with social prescribers. Patient and Public Involvement (PPI) representatives had input throughout all development work. Findings from these activities were triangulated to inform the development of a prototype intervention which was tested in a repeated-measures, mixed methods proof-of-concept study called De-Stress *Pain*.

The De-Stress *Pain* intervention offered 4-6 social prescriber sessions over 12 weeks, and access to a study website. 16 participants were recruited from general practices, and 4 social prescribers were recruited from Primary Care Networks (PCNs) and charitable organisations. Both quantitative measures and qualitative interviews were completed by participants at baseline and 12 weeks post-baseline. Quantitative measures included questions about mood (DAPOS, WEMWBS, 4DSQ), pain chronicity, pain intensity and musculoskeletal health (MSK-HQ). Semi-structured interviews with participants explored the participants’ context, their experiences of pain related distress, and their experiences of the intervention. Social prescribers were interviewed to explore their experiences of delivering the intervention. Interviews were analysed using thematic analysis.

**Results:** The De-Stress *Pain* intervention was acceptable both to patients and to social prescribers. Some participants experienced positive changes such as improved mood, increased hope and increased activity. The social prescribers (“De-Stress Coaches”) provided accountability and supported motivation. All measures of mood showed improvement. Limited time and money were identified as barriers to engagement, along with participants holding the view that increasing pleasurable activities was indulgent. Some participants were already socially engaged and busy at the point of entering the study.

**Conclusions:** We have confirmed the intervention is needed, acceptable and welcomed by people with pain. Social prescribers found the intervention acceptable to deliver. We identified the barriers that need to be addressed in future versions of the intervention.

## Background

The latest comprehensive evidence review of musculoskeletal health in the UK(2) paints a concerning picture: over 20 million people report a persistent musculoskeletal (MSK) condition. It is the third largest area of NHS spend and one of the biggest contributors to years lived with disability (YLDs)(2). Pain is the primary reported MSK symptom, affecting 74% of people with MSK conditions. Of these, 42% suffer from back pain, the leading cause of years lived with disability in the UK(2). MSK conditions are the third most common reason for working days lost in the UK(2). As of March 2023, 995,000, people in the UK are economically inactive due to problems or disabilities connected to their back or neck(3). Persistent MSK pain is predominantly managed in primary care with people living with MSK accounting for 1 in 7 GP consultations(2).

Depression is four times more common in people living with long term pain conditions(2). People living with MSK pain have reported they often struggle to live well(4). Plans are difficult to make due to uncertainty, physical demands, and fatigue; relationships can become strained, and work is challenging. People have reported feeling like a burden, live with guilt and low mood, and withdraw from many of their preferred activities(5, 6). They also report frustration and disillusion with the support provided by their doctors, both for their pain and their low mood(6). Doctors in turn have told us that it can be difficult to help these patients(6). Guidelines recommend a trial of tricyclic antidepressants for chronic pain(7), and for more severe depression(8), but these are not always helpful or acceptable to patients(9). In addition, treatment services such as pain management, and NHS Talking Therapies (previously known as IAPT – Improving Access to Psychological Therapies’) offering predominantly cognitive behavioural therapy and behavioural activation, can have long waiting lists(10). Even after being discharged from NHS Talking Therapy services, or completing pain management courses, which typically emphasise self-management, patients find themselves in a support void(11).

Having to withdraw from cherished activities because of pain results in distress and can impact on self-identity(12). Often this can be accompanied with considerable efforts to cure or reduce the pain. Pincus and Morley (2001) reviewed evidence of cognitive bias in people living with pain and postulated that the enmeshment between self and pain is at the root of pain-related distress and depression(12). As the concept of self becomes closely associated with the concept of pain, other thoughts and memories are reduced. This is compounded by a reduction in enjoyable activities(12). Our research focuses on reversing this enmeshment.

A combination of validated, evidence-based psychological theories refined our approach. One approach to reduce enmeshment and increase wellbeing is Acceptance and Commitment Therapy (ACT), based on the premise of living life to the full amidst the pain(13). A recent overview of systematic reviews of ACT interventions suggests that it is effective at reducing depression and increasing function and wellbeing(14), showing promise at improving wellbeing and reducing distress in the long run.

ACT is an action-oriented approach requiring patients to commit to making changes to bring them closer to their desired outcomes(15). ACT is particularly appropriate in the context of pain related distress because it is based on the underlying assumptions that fighting unavoidable pain leads to exhaustion and misery and doing things you enjoy with people who matter to you will improve wellbeing and mood. Selecting activities that are fundamentally aligned with an individual’s values is key.

We were also guided by self-determination theory(16), which suggests that self-motivation can be improved by fulfilling individual’s needs for autonomy, competence and relatedness. I.e., to achieve psychological growth, one needs to feel: in control of behaviours and goals (autonomy); confident one has the skills needed for success (competence); and a sense of belonging and attachment to others (relatedness or connection). A recent systematic review with meta-analysis suggests that interventions based on self-determination theory are more likely to result in improvements to psychological and physical well-being than usual care, and of importance, result in changes in behaviour(17).

Finally, we were interested in incorporating principles from behavioural activation in our approach. Behavioural activation is considered a parsimonious intervention, using the least complex but acceptable theoretically derived treatment(18). Such interventions have been found to be effective for people living with depression, where typically they encourage people to reconnect with positive environments through activity scheduling(19).

In recognising the need to reduce pain-related distress in people living with chronic MSK pain, our aim was to develop a patient-centred intervention to help this population. Intervention development was guided by the person-based approach (PBA)(1), which is an established framework providing a clear process to ensure interventions are grounded in the perspectives and psychosocial context of the target user group(20). We conducted a proof-of-concept study to test the intervention’s acceptability.

## Intervention planning methods

The PBA recommends qualitative research with the target user group to gain a rich understanding of their key beliefs, the challenges they face, the factors that may facilitate change and the barriers that may hinder the effectiveness of an intervention. Following the PBA, resulting interventions are influenced by target users’ needs and preferences, improving acceptability and user engagement(1). We commenced our target user studies with interviews with GPs and people with pain, and used the results to guide our next activity, which was a stakeholder discussion with social prescribers. We discussed our findings with the study PPIE group and worked with them to develop a lay-person model of change. We used the findings from these activities to create key guiding principles and a logic model (see Figure 1 below).

**Figure 1:** Flow chart to show intervention planning and development process.

### Qualitative study with GPs and people with pain

In work leading to this study, published elsewhere(6), semi-structured interviews were conducted with people with persistent pain (n=21) and General Practitioners (n=21) to explore perspectives and understanding of primary care management of people with persistent MSK and distress. The findings from these interviews were discussed with our PPIE group, and provide the following foundations for the De-Stress *Pain* intervention (Table 1):

**Table 1:** Foundations of the intervention resulting from interview findings and informed by PPIE.

| Findings | Supporting data | Intervention implications |
| --- | --- | --- |
| General Practitioners felt 'stuck' when dealing with pain-related distress, and patients felt that there was little that could be offered to them by their GPs. | GP 6: *"As GPs we want to help them but we're struggling because there's no resources. If you know the NICE guidelines which came last year about chronic pain management, clearly they said no to any analgesia for that matter. So again, we've limited resources to manage these kind of patients."*<br>Person with pain (P5): *"Well there was nothing I could do really, you know, nothing, I couldn't do anything to improve it, you know, the only thing that flagged it was the painkillers."* | • Delivery outside of medical setting.<br>• Not delivered by GPs. |
| GPs recognise the difference between depression and pain related distress, and are therefore likely to be on board with the objectives of the intervention | GP 5: *"Distress and depression, well I think... distress is when I think you know there's something that's happened specifically and the distress or the emotional upset is related directly to that thing."* | • Recruitment through primary care is viable. |
| GPs considered that social prescribers are ideally placed to encourage social reactivation, and can give more time than GPs. | GP 9: *"The other thing we have is a social prescriber now within our practice and that's really good. We haven't got painkillers for these people. They don't need surgery and they don't need doctors. They need a life, basically...A lot of these people are very deconditioned and so I'm keen to get people exercising but I recognise that sending people to the gym is not really going to do it. So walking groups, gardening groups and that kind of thing or anything that gets patients socially engaged with their community."*<br>GP 12: *"Using social prescribers is good* | • Delivery by social prescribers would fit within NHS systems and address intervention principles. |
|  | *because they have more time per client than the GP, with the best will in the world we can't spend an awful lot of time with any particular patient about emotional distress. Although obviously it's our role to differentiate between that and true depression."* |  |
| There was strong agreement amongst patients that a person delivering one-to-one contact was essential. The mode of contact mattered less. | Person with pain (P12): *"If you just knew there was one person who was your case worker and they would deal with the GP, and the consultant, and they would be the person that says, 'Right, to get from here, what we need to do is just get it working.' I think that would be beneficial...you just want one person, so you wouldn't have to repeat the same thing 15 times"* | • Participants need 1:1 contact with a person. |
| There was support for online information in addition to person-based delivery, but patients emphasised the information must be accessible to all. | Person with pain (P8): "Yeah that's [both face-to-face and online] probably better. You know, you actually see somebody and speak to them, but do stuff in your own time as well. But you've got somebody explaining or just following up and just making sure that you're doing what you're supposed to." | • Create a simple, user-friendly website, ensure use of lay language, images and ease of navigation. |
| Patients and PPIE told us that the most distressing aspects of living with pain were inability to do the things you used to enjoy, reduced social contact, and difficulty making plans. | Person with pain (P3): "it's affected my well-being...not being able to do what I want to do. I used to be a runner and I can't run anymore and that really has devastated me."<br>Person with pain (P8): "So normally trying to decide what time of day I'm going to do certain things knowing that I'm going to be in pain afterwards is difficult. You know and having to plan how you're going to do everything, what you're going to leave out, | • Base the intervention on enjoyable activities.<br>• Allow for opportunities to carry out some activities with other people.<br>• Allow flexibility around approaching activities (no schedule, no use of |
|  | what doesn't need doing, do I use my energy. It's not even energy, but do I use my good part of the day to do something that I enjoy or something that I need to do? Yeah because doing the two isn't easy." | the word 'should'). |
| Addressing pain related distress will take more than one consultation and continuity of care is important | GP 19: "I would say that's the first and the most important thing and having continuity of care with the same doctor or the same clinician and not seeing a different person every time. It should be someone who really gets to know the patient and understands their point of view and the patient then, over time, gets to know and trust that doctor. I don't think you can do it in one consultation." | • Offer more than a single session, and ensure sessions are with the same person. |
| Moving the focus away from pain is helpful | Person with pain (P13): "I suppose I've actually covered it myself really because it's a sense of wellbeing. Things that I used to enjoy doing... I did used to be in a choir and that helped too... it's something that takes you to a different place and stops you focusing on your pain. As I say, it will never take your pain away, obviously. Let's say you go and sing in a choir for an hour, that's an hour that you're not thinking about your pain... That puts the pain a little bit out of my mind." | • Don't focus on pain. |
| Credibility is important | Person with pain (P04): "I think professionally doctors or people like you or research people, because you've got such a mind to do what you do, I think you can trust in what you're saying...I would feel so much trust in anything that you would tell me because I know that you're going to go to the right sources for things. So I think in that aspect professionals are invaluable because like arthritis research | • Intervention needs to be delivered by experts in social reactivation. |
|  | I'll go to them because I know they're up to the minute with all the research that's coming out and I can feel confident in what I'm reading. So I think yeah, a person who is specialising in something and you know that that's their passion and that's what they do, you can trust in it. You can feel completely relaxed. I know I can go to this and it will tell me what I need to do or hear." |  |

### Stakeholder discussion with social prescribers

Our exploration of the needs of people with pain suggested social prescribers might be suitable to deliver an intervention for this population. To further explore this idea, we held an online stakeholder discussion with 9 social prescribers. The social prescribers were part of a charitable organisation working across The Midlands in the UK. To facilitate our understanding, we asked questions about what they do in their role, how many sessions they typically offer, whether they conduct these face-to-face or by telephone, whether they currently see people with pain-related distress, and the challenges they face. We then explored their thoughts about our intervention specifically for pain-related distress being delivered by a social prescriber and whether they felt this was something they could deliver, and the challenges that they might face. Our main findings are presented below in Table 2. For reasons out of our control this discussion could not be recorded and therefore we cannot cite direct quotes.

**Table 2.** Stakeholder discussion suggestions and implications for the intervention.

| Suggestions | Implications for the intervention |
| --- | --- |
| Social prescribing is tailored according to individual needs. | Chronic pain varies. Our participants will each have different needs. The social prescribers' ability to tailor to these needs will be beneficial. |
| Social prescribers try to give clients ownership | Participants will be equipped to continue |
| over decisions and encourage independence. | changes after intervention has finished. |
| Social prescribers can give more time to the clients compared to other health care professionals. | Corroborates findings from our 'exploration of population needs' where GPs explained these patients need more time with a professional. |
| The social prescribers in this group did not offer a set number of sessions. The number of sessions depends on the client's situation and needs. | For the purposes of research, a maximum number of sessions over a set period of time will be necessary. |
| Social prescriber sessions are offered either face-to-face or by telephone, and this flexibility is valuable to their clients. | Our intervention should offer the flexibility of telephone or face-to-face. |
| Social prescribers sometimes see people experiencing pain-related distress, but feel they are more commonly referred to Health and Wellbeing Coaches where they focus more on the pain than their mood and life outside of the pain. | People with pain-related distress are often encouraged to address the pain and believe mood will improve once the pain improves. The intervention needs to be different and focus on mood. |
| Social prescribers identified client transport (cost and availability) as a barrier. | Important for the intervention to offer 'at home' options and consider ways of getting out of the home despite transport issues. |
| Social prescribers believe an intervention focussing on mood for those with chronic pain would be valuable. | Social prescribers on board with the concept of the intervention. |
| Social prescribers feel comfortable and qualified to deliver an intervention for those with pain-related distress and believe it would be very similar to the work they do daily. | Social prescribers will need minimal training for the intervention. |

## Intervention development

Informed by the PBA framework we used the findings from the above exploratory work to create a lay-person model of change, Key Guiding Principles, and a logic model.

### Lay-person’s change model developed with PPIE

We presented our findings from the interviews with people with pain and GPs, and the discussion with social prescribers to our PPIE group. Together we developed a model to demonstrate how they see change occurring within De-Stress Pain (Figure 2).

**Figure 2:** Lay-person’s model of change.

### Key Guiding Principles

Key Guiding Principles summarise the design objectives and how they will be achieved. These principles focus decision making throughout the development phase(20), see Table 3 below.

**Table 3.** Key Guiding Principles.

| User context | Key design objective | Intervention |
| --- | --- | --- |
| People with pain suffer low mood. | Ensure users understand the difference between clinical depression and pain related distress. | Provide clear explanation about the difference between clinical depression and pain related distress. |
| People with pain have lost their identity. | Help users re-find their sense of self. | Use the video 'Pain and me'(21) to communicate the message 'you are bigger than the pain'. Encourage users to compartmentalize pain so it is a PART of them, rather than ALL of them. Shift focus onto the parts not impacted by pain. |
| People with pain have stopped engaging with the things they love. | Encourage social reactivation. | Offer suggestions of activities that might be possible. |
| People with pain focus on pain. | Clear rationale about why shifting focus onto living well will be beneficial. | Focus on how an individual's life/psychological wellbeing can be improved. |

### Refining the intervention with psychological theory

The main premises of Acceptance Commitment Therapy (ACT) and Self-Determination Theory were considered when refining intervention design. Table 4 below describes the implications of these theories for our intervention.

**Table 4:** Psychological theory and our intervention.

| **Theory** | **Premise** | **Implications for intervention** |
| --- | --- | --- |
| Acceptance<br>Commitment<br>Therapy (ACT) | ACT encourages acceptance of feelings and emotions about a certain situation (in this case chronic MSK pain) and aims to move the focus to ways in which one can move forward. | Intervention should not focus on pain. Instead, it should focus on positive, pleasurable activities. |
| Self-determination theory | Self-motivation can be improved by fulfilling individual's needs for autonomy, competence and relatedness. I.e. one needs to feel in control of behaviours and goals (autonomy); confident one has the skills needed for success (competence); and a sense of belonging and attachment to others (relatedness or connection). | Participants need to be encouraged to make a personalised plan based on their preferences, allowing them to feel in control. Participants need to be guided in making their plan to ensure it is possible (e.g. they have the equipment needed, or can travel to the activity). Participants need to be encouraged to include activities that are social/with other people to improve feelings of connection. |

### Logic model

As recommended by the MRC complex intervention guidelines(22), we developed a logic model to outline the hypothesised causal mechanisms involved in bringing about change. Figure 3 below shows the logic model for how our intervention would bring about change in people with pain-related distress according to our development work.

**Figure 3:** Logic Model.

### The Intervention

The intervention planning and development phases resulted in a prototype intervention. The De-Stress Pain intervention was designed to offer of 4-6 sessions with a social prescriber (named ‘De-Stress Coaches’ for the purposes of the intervention) over 12 weeks. Drawing upon information obtained in our background discussions with social prescribers we decided 4-6 sessions was the most appropriate amount of contact to allow for participants to create and adjust personal plans where needed. The De-Stress coaches received 3 hours training with the research team and a manual to guide them through sessions. Participants were offered access to a study website which included modules about self-kindness, the rationale behind increasing pleasant activities, and provided suggestions for local activities that participants might like to try. The list of suggested local activities was developed with PPIE and included walking groups, painting, pottery, swimming, online film clubs and book clubs. The De-Stress Coach sessions aimed to:

- Help participants identify accessible pleasurable activities and create a plan to carry them through.
- Identify ways to adapt activities when needed (for example, when experiencing pain flare-up).
- Deflect conversations about pain and encourage participants to focus their energy on positive changes they can make.
- Support participants if their plans need changing.

Figure 4 below outlines the intervention sessions.

**Figure 4:** The intervention.

The intervention was tested for acceptability in a proof-of-concept study.

## Methods for proof-of-concept study

### Aim

To test the acceptability and proof of concept of an intervention to reduce pain-related distress in people with chronic MSK pain.

### Design

A repeated measures mixed-methods proof of concept study consisting of a quantitative survey and qualitative interviews. Both quantitative measures and qualitative interviews were completed at baseline and 12 weeks post-baseline.

### Recruitment

De-Stress Coaches: Social Prescribers were recruited as De-Stress coaches via Clinical Research Network mailouts, and the BNSSG (Bristol, North Somerset, and South Gloucestershire) Training HuB. Participants were recruited between 01/09/2023 and 31/12/2023.

Participants: People with persistent pain were identified through GP practices who searched patient databases using relevant SNOMED codes. 593 potential participants were sent an invitation letter with details about the study.

### Inclusion criteria

- Chronic MSK Pain (MSK pain ongoing for more than 3 months e.g. knee, hip, back pain, fibromyalgia)
- Aged over 18
- Ability to complete questionnaire and interview in English
- Ability to provide informed consent

### Sample

Seventeen participants consented (via an online consent form) to the De-Stress Pain intervention, along with 4 De-Stress Coaches who led the coaching sessions. Sixteen participants went on to have sessions with a De-Stress Coach and provided qualitative data, 12 provided quantitative data at 12 weeks.

### Survey study methods

#### Quantitative measures

All questionnaires were completed online. Potential participants completed a screening questionnaire before proceeding to the baseline questionnaire. The screening questionnaire ensured they met the eligibility criteria described above, and that they were not experiencing suicidal thoughts. If a potential participant was not eligible they were informed immediately. Those who passed screening were given instant access to the baseline questionnaire. A follow-up questionnaire was issued at 12 weeks post baseline. Table 5 below describes the measures and the timepoint(s) they were completed:

**Table 5:** Measures included in the survey.

| Variable | Measure | Baseline or 12 week follow-up? |
| --- | --- | --- |
| Clinical and demographic characteristics | Type of musculoskeletal pain, age, gender, education, ethnicity, employment status. | Baseline only |
| Mood | Single item to distinguish distress due to pain, due to other things, or both. | Baseline only |
|  | Single item rating distress 0-10 over last 2 weeks. | Baseline and follow-up |
|  | DAPOS, 11-item scale to assess depression, anxiety and positive outlook (23). | Baseline and follow-up |
|  | WEMWBS, 14-item scale to assess wellbeing(24). | Baseline and follow-up |
|  | 4DSQ, four subscales measuring: distress (16 items), depression (6 items), anxiety (12 items) and somatization (16 items)(25). | Baseline and follow-up |
| Pain chronicity | Single item question to assess chronicity (how long since no pain for a month)(26). | Baseline only |
|  | Single item asking number of troublesome days in pain in the last 4 weeks(27). | Baseline and follow-up |
| Pain intensity | Numerical rating scales rating worst, least, average, and current pain, and coping(28). | Baseline and follow-up |
| Musculoskeletal health | MSK-HQ, 15-item scale to assess quality of life and pain interference in patients with MSK pain(29). | Baseline and follow-up |

### Qualitative methods

#### Interviews with participants

Semi-structured interviews with participants were conducted at two timepoints (baseline and 12 weeks post baseline) using an iterative topic guide.

- Baseline interview: Conducted after the participant completed the baseline questionnaire, and before they started the intervention. This interview gathered insight into the participant’s context, their experiences of pain-related distress, why they wanted to be part of the study and their expectations of participation.
- Follow-up interview: Conducted after the participant completed all sessions with their De-Stress Coach and the quantitative questionnaire. The interview explored their experiences of the intervention, any changes they had made since taking part, and any barriers or facilitators they came encountered.

#### Interviews with De-Stress Coaches

Semi-structured interviews were conducted with the De-Stress Coaches at the end of their involvement in the study. These interviews explored overall views of providing support as part of the study, perceptions of participant experiences, the relationships they built with participants, their views about the training and manual provided as part of the study, and their thoughts on limitations and future research.

#### Data analysis

Interviews were transcribed verbatim, imported into NVivo and analysed by authors HB and SH using thematic analysis (Braun & Clarke 2006).

## Results

4 De-Stress Coaches were recruited working across 3 primary care practices. 17 participants consented to the study and completed the baseline questionnaire, and 12 participants completed the follow-up questionnaire. 16 participants participated in the baseline interview, and 11 completed the follow-up interview.

### Participant Characteristics

Recruited participants were mostly white (n=16), female (n=12), and ages ranged 24-79, with a mean age of 53.2. The employment status of participants was mixed, full-time (n=7), part-time (n=2), retired (n=5), unemployed (n=3).

### Survey results

Table 6 below shows the mean scores and standard deviations for each of the measures at baseline and 12-week follow-up. The study is not powered to assess significance and meaningful conclusions cannot be drawn from these results.

**Table 6:** Mean scores and standard deviations at baseline and 12-week follow-up.

| Scale | Direction of improvement | Baseline (n=17)<br>Mean (SD) | Follow-up (n=12)<br>Mean (SD) |
| --- | --- | --- | --- |
| Pain intensity | lower | 5.18 (1.33) | 5.58 (1.62) |
| Pain coping | higher | 4.94 (1.71) | 6.08 (1.88) |
| Distress due to pain | lower | 6.60 (1.62) | 5.25 (2.09) |
| DAPOS – depression | lower | 13.71 (4.04) | 10.83 (4.32) |
| DAPOS – anxiety | lower | 8.24 (3.09) | 6.75 (2.80) |
| DAPOS – positive outlook | higher | 8.71 (2.59) | 9.67 (2.06) |
| WEMWBS (wellbeing) | higher | 38.59 (8.38) | 42.92 (7.95) |
| MSK-HQ | lower | 31.71 (7.10) | 27.58 (8.49) |
| 4DSQ depression | lower | 4.24 (3.77) | 3.50 (3.87) |
| 4DSQ distress | lower | 20.05 (7.60) | 16.5 (8.21) |
| 4DSQ somatic | lower | 15.64 (6.72) | 13.50 (6.80) |
| 4DSQ anxiety | lower | 7.82 (6.51) | 7.25 (6.80) |

### Qualitative findings

At baseline, participants displayed a strong belief in the relationship between pain and mood. They understood and supported the concept of pain-related distress and that it is different from clinical depression. Prior to the intervention participants described feelings of frustration, irritability, anxiety and low mood. Pain-related mood impacted their relationships, many withdrew from activities and reduced social interactions. When presented with the intervention participants described feelings of enthusiasm, hopefulness and appreciated a refreshing, new approach.

Five main themes were developed from analysis of follow-up data. These themes are shown in Figure 5 and expanded in the text below.

**Figure 5:** Key themes.

#### Participants reported benefit from the intervention

The majority of participants who engaged with the intervention reported positive changes due to taking part in the intervention, such as, increased activity, improved mood, increased hope and a new understanding of the benefits of doing things they enjoy. One participant described feeing stronger, healthier and happier:

> *“I just feel stronger in myself… healthier in myself…I’m just happier as well, I’d like to say. I’m more open…” (P015, M)*

Before the intervention, participants described feeling frustrated because they were unable to do the things they enjoy due to pain. The De-Stress coaches were able to discuss and address the barriers of these activities to find acceptable solutions. One participant described how a De-Stress Coaching session helped her see how alterations to the activities she enjoys could make them manageable, and in turn increased her feelings of hope:

> *“I do feel like I came out of that session with a lot more hope that actually there are so many things that I could alter so that I can do more. I hadn’t thought of the alterations she’d suggested, so it gave me a bit of an, oh, there is something I can do about this.” (P004, F)*

Although some participants described knowing that doing things they enjoy would be good for them, taking part in the intervention spurred a realisation that it is important to make time to do these things. Participation gave them permission to prioritise their wellbeing, and, when they started to re-engage, the importance of these activities was reinforced:

> *“I’m also realising that I do need to do those things [pleasurable activities], and if I don’t, it’s just long-term issues… I’m recognising that I’ve got to put that effort in because I know it makes a difference when I do.” (P009, F)*

Once participants started participating in activities they enjoy, they reported that their attention was focussed more positively than before, and participants described a clear shift in mental wellbeing:

> *“Even though the pain still feels the same, and it still hurts the same, because my mind’s thinking about other things, it doesn’t feel like it’s hurting as much, so I haven’t had as many painkillers or anything, so that’s better. And my head feels clearer.” (P016, F)*

#### The De-Stress Coaches supported motivation

Participants explained having someone to ‘report back to’ helped them keep on track. The accountability this provided meant participants were more likely to push themselves into carrying through with their plans:

> *“Accountability to actually make yourself do it because we all know that these things are a really good idea, but actually having one-to-one support and saying, so what did you find? Have you done that? Come back to me and let me know how you found it. It’s like, okay, I’m going to have to do it then, aren’t I?” (P009, F)*

Receiving the support from a real person (rather than online) was valued, and was reported to positively impact on engagement:

> *“I think it probably did help having an actual person. I think if I’d been given a website, I would have looked at it and gone, ‘that might have been a good idea, but I can’t really be bothered to do it.’ Then I knew I was going to speak to her and she was going to say, ‘oh, I’ve looked at all these things for you, what do you think?’ I wasn’t going to go, ‘well, I’ve not looked at anything you’ve sent me!’” (P004, F)*

Participants described the De-Stress Coaches with words such as ‘invested’, ‘understanding’ and ‘helpful’, and valued the rapport and relationships they formed, which in turn enhanced engagement.

#### Participant budget and availability restricted activity choice

Many participants were restricted in the activities they chose due to money, and some reported choosing activities that were free, rather than the activities they find pleasurable because of their financial constraints:

> *“it was the cost of things as well. That’s the reason, really, that I went with the up-to-six-months gym membership thing that that was funded, whereas the pottery classes were going to be £30 a time.” (P005, F)*

It was suggested that participant budget was considered before activities were suggested:

> *“the sessions need to focus on people with different income levels or looking into funding for things before they get offered to you.” (P005, F)*

Another barrier to engagement was time; participants who were busy, for example with work or existing activities, found it difficult to engage in appointments with the De-Stress Coach and adding activities to their schedule:

> *“The main thing is it’s just been tough organising, trying to do an appointment…whether I can make it a certain time. I’ve been really busy with work.” (P007, F)*

For those who worked shifts it was difficult to commit to a regular activity at a set time, and participants stressed the need for flexibility in the activities they chose.

#### Some participants were already socially active

Participant interviews pre-intervention revealed that some agreed to take part because they wanted to help and facilitate understanding of pain-related distress, as opposed to feeling the intervention may benefit them personally. The De-Stress Coaches questioned whether these participants were the population most in need of the intervention, as many were already socially engaged and active:

> *“I think it almost did feel a little bit like the patients that I was talking to were maybe a little bit too engaged in that they already have quite a lot of stuff going on…and slotting in other stuff did feel like it was a little bit difficult sometimes.” (SP002)*

With these participants De-Stress Coaches reflected that it was a struggle to know how to add anything:

> *“She talked about things she liked, and she liked a lot of things…she was the person that just tried to get on with life and try to put the pain to one side. Yes, she was involved in lots of local groups, committees. It was challenging in that I didn’t really know what else I could offer.” (SP001)*

Despite this, the De-Stress Coaches reported finding ways to help such participants, for example, by supporting them to manage their time and prioritise the things that were most important and likely to benefit their wellbeing.

#### Pleasurable activities are an indulgence

Some participants felt their responsibilities (for example, work, or caring for family) needed to take priority and viewed participating in pleasurable activities for themselves as an indulgence:

> *“I know how that sounds, but it’s not a priority, my well-being. Even washing my hair is not a priority in comparison to making sure the kids have got their school uniform, have done their homework… It’s really hard. I mean, if it was a doctor’s appointment, that’s quite a priority, you know, but because it’s something just for me to try, it’s an indulgence.” (P007, F)*

This formed a barrier to engagement. Furthermore, understanding that doing pleasurable things is important and beneficial to wellbeing facilitated engagement. Participants who consciously made the decision to prioritise their wellbeing found the process beneficial:

> “It’s been quite selfish in a way that I’ve had to look after myself now for a bit. My kids are 20 and 16, but it’s been quite nice for me to look after me for once… I’ve had to look after myself this time.” (P015, M)

## Discussion

### Summary of findings

The majority of participants in this proof-of-concept study found the De-Stress Pain intervention acceptable, valuable and welcomed this new approach. Some reported positive changes due to participating in the intervention, such as, improved mood, increased hope, a more positive outlook, and increased activity. Accountability is important for motivation, and this needs to be in the form of contact with a real person. Viewing an increase in pleasurable activities as an indulgence, or a luxury was identified as a barrier to engagement. Other barriers included participant availability due to other commitments such as work and family, and participant budget. Some participants recruited to this proof-of-concept study were already busy with social and pleasurable activities, suggesting we did not reach our target population.

### Findings in relation to previous literature

Addressing pain in interventions for people with chronic MSK pain, for example, by including pain management techniques or targeted exercise, is commonplace(30–34). However, evidence from trials of the most effective interventions, such as physical activity or CBT, suggest they yield at best small-effects, and that these might not last in the long term(35). There is also evidence that after attending pain management courses, some patients find themselves in a support void, in which they struggle to adapt the principles they learnt to their life outside of the care content(11). The principles of the De-Stress Pain intervention are supported by research on self-schema in people living with chronic pain. In a review and integration of evidence on cognitive biases, Pincus and Morley (2001) suggested that enmeshment between pain-distress and self was at the core of cognitive changes that perpetuated low mood(12). Defusing self from pain is the primary target of our new intervention. Although it is based on ACT principles, such as experiential learning in the here and now, there are several important differences between the new intervention and traditionally delivered ACT for pain. Typically, patients will attend sessions in a clinical setting, and sessions will include attempts to foster acceptance of living with pain through creative hopelessness - the realisation of the cost and limitations of attempts to control or cure pain(13). Such discussions can be uncomfortable but are considered necessary to accept and live better with pain. The De-Stress Pain intervention by-passes this stage completely and focuses instead on ‘doing’. The hypothesis is that diffusion between pain and self will happen automatically as self is enriched with rewarding activities. Our novel approach purposely deflected focus and conversations about pain, and instead, focussed energy on positive, pleasurable, attainable changes. The results show promise for this approach.

A previous review of 15 social prescribing interventions reported methodological shortcomings such as; a lack of validated and standardised reporting measures, short follow-up durations, and a failure to consider potential confounding factors(36). Unlike the programmes included in this review, the De-Stress Pain intervention is strengthened using validated measuring tools. As a small proof-of-concept study our follow-up time was relatively short (12 weeks), and we were unable to account for confounding factors, however, these factors will be addressed in future versions of the intervention. We note that De-Stress Pain offers social prescribers training in the underlying principles of the interventions, and in improving delivery through a support manual.

### Strengths and limitations

Persistent MSK pain and the impact it has on a person varies widely. Using the PBA to guide development has resulted in a core product based on a rigorous transparent process, with clear guiding principles that can be adapted and updated according to the specific needs of the population, meaning the intervention can be adapted depending on factors such as participant mobility, pain location, ability to travel and budget. De-Stress Pain was developed iteratively and inductively. PPIE were involved throughout the planning and development processes ensuring our decisions were relevant to people with MSK pain.

Due to the small sample size, and because over 25% of our sample did not provide data for the 12-week follow-up, the quantitative data has limitations. As a small proof-of-concept study we acknowledge the limited generalisability of our findings to the wider population, or to people with pain i.e., the sample was small, located in two geographical locations and the recruitment strategy did not attract an ethnically diverse sample. However, our sample was adequate to assess acceptability before rolling out to a larger sample.

Qualitative data analysis suggested some of our participants were not those most in need of De-Stress Pain and were already leading full lives. Our recruitment methods did not exclude those who wished to take part in the study purely to ‘help others’, rather than feeling the intervention might be something they would benefit from, and this may explain the limitation of our sample.

### Implications for future research and conclusions

As discussed in the qualitative findings, some participants felt increasing pleasurable activities was an indulgence, which was a barrier to engagement. Future versions of De-Stress Pain need to address this barrier by clearly addressing the importance of looking after one’s own wellbeing in the information given to potential participants, and the early stages of the intervention. The structure of De-Stress Pain appears to work for social prescribers and people with MSK pain. This includes the combined in-person and online approach. Future research should monitor which aspects are most used and include analysis of acceptability in a large sample.

Future versions need to establish the financial and time restrictions of the participants at the first coaching session, and only offer or suggest activities that are possible for the individual. Where possible, there needs to be flexibility in the timing of De-Stress Coaching sessions to improve accessibility, for example, out of usual working hours. Recruitment methods for future research should strive to recruit those most in need of the intervention, for example, those less engaged and in need of social reactivation. Future recruitment strategies must aim to recruit an ethnically diverse sample to improve generalisability.

This is a novel approach to supporting people living with pain-related distress. The results show promise for future, larger trials.

## Data Availability

All data produced in the present study are available upon reasonable request to the authors

## Ethical approval and consent to participate

Ethical approval for this study was granted by London – Bloomsbury Research Ethics Committee (reference: 23/LO/0676). All participants provided informed consent to participate, and for publication of anonymised data.

## Contribution of authors

SH: Design of the work, analysis, interpretation of data, drafting the article.

TP: Conception and design of the work, substantially revising the article.

AG: Conception and design of the work, contributions to the article.

CCG: Conception and design of the work, contributions to the article.

BS: Design of the work, analysis.

PL: Conception and design of the work, contributions to the article.

MM: Conception and design of the work.

HB: Design of the work, interpretation of data, substantially revising the article.

